# Metabolomic and transcriptomic signature in Kabuki syndrome

**DOI:** 10.1101/2025.04.30.25326738

**Authors:** Youngsook Lucy Jung, Christina Hung, Jaejoon Choi, Eunjung A Lee, Olaf Bodamer

**Author notes:** Correspondence (O.B.).

## Abstract

Kabuki Syndrome (KS) is a rare multisystem disorder with a variable clinical phenotype. The majority of KS cases are caused by dominant loss-of-function mutations in two genes, *KMT2D* (lysine methyltransferase 2D, KS1) and *KDM6A* (lysine demethylase 6A, KS2). Both *KMT2D* and *KDM6A* play a critical role in chromatin accessibility, which is essential for developmental processes and differentiation. In a previous study, we reported that *KMT2D* mutations could lead to increased enhancer activity in genes related to metabolomic pathways in KS1. Early detection of KS is crucial in order to offer improved treatment options. To uncover new biomarkers that could facilitate early detection and to inform clinical trial readiness, we conducted a study in which we collected and analyzed plasma and urine metabolites from 40 KS patients with pathogenic mutations in either *KMT2D* or *KDM6A* and 12 healthy controls. We employed an untargeted approach using Liquid Chromatography with tandem Mass Spectrometry (LC-MS/MS). Additionally, we profiled gene expression in the most KS patients and controls. Our analysis revealed > 100 significantly altered metabolites between KS patients and controls, with these metabolites being clustered based on genotypes. Importantly, we identified N2, N2-dimethylguanosine emerging as one of the top candidates in both KS1 and KS2 patients. We utilized machine learning classifiers and identified the most crucial metabolites. Using this trained model, we achieved a high level of discrimination between the KS data and controls. Furthermore, pathway analysis revealed several disrupted pathways, including the pyrimidine metabolism pathway, which are associated with the significantly altered in both metabolome and transcriptome in KS. Distinctive metabolites identified in KS can effectively serve as discriminative biomarkers. Our findings provide valuable insights into the metabolic dysregulation underlying KS and highlight potential targets for further investigation and therapeutic interventions.

## INTRODUCTION

Kabuki syndrome (KS) is a rare congenital disorder. The various symptoms with KS are manifested in multi-organs, such as abnormalities in cranial facial features, intellectual disability, hypotonia, immune functions, development delay, hearing loss, and heart and kidney malformation(Niikawa et al. 1981; Kuroki et al. 1981; Adam et al. 2019). This condition affects approximately 1 in 32,000 individuals and currently lacks approved therapies. KS conditions impose significant burdens on both patients and caregivers(Adam et al. 2019; Theodore-Oklota et al. 2022). For example, patients with KS encounter challenges in medical conditions, learning delays and socio-emotional issues. KS is largely heterogeneous in nature. Patients with KS exhibit varying degrees of impairments involving different organs. This heterogeneous nature in KS makes early screening and diagnosis of KS patients difficult.

KS is primarily caused by de novo mutations in two genes, *KMT2D* (lysine methyltransferase 2D, KS1, MIM: 147920) and *KDM6A* (lysine demethylase 6A, KS2, MIM: 300867). Both of them play pivotal roles in modulating chromatin accessibility(Froimchuk, Jang, and Ge 2017; Lavery et al. 2020). *KMT2D* is responsible for adding methyl groups to H3K4, while KDM6A removes methyl groups from H3K27. Although the exact etiology of KS is unknown, previous studies suggest that KS is associated with various metabolic abnormalities. *KMT2D* regulates hepatic metabolism, therefore its dysregulation resulting in the increased levels of bile acid (Timothy et al. 2019). A mouse model of KS exhibited metabolomic alterations (Benjamin et al. 2017). We previously showed that signals at promoter-proximal enhancers of metabolism associated genes are altered in KS compared to controls (Jung et al. 2023). These findings suggest that mutations in *KMT2D* or *KDM6A* would potentially alter the metabolomic landscape in KS.

Untargeted metabolomics has emerged as a promising tool in screening for underlying metabolic changes, therefore exploring possible new biomarkers for disease conditions. Here, we profiled 40 patients with KS, for whom we profiled untargeted plasma and urine metabolites as well as gene expression. We identified KS-specific metabolomic alterations that hold potential for biomarkers and clinical trial readiness in KS.

## RESULTS

### Characteristics of cohort and datasets

Urine and plasma samples were collected from a total of 52 subjects enrolled in this study: 32 individuals with KS1 due to pathogenic mutations in *KMT2D* (males 31.3 %, mean age 9.9 years, age range: 0.9 - 33.2 years), 8 individuals with KS2 due to pathogenic mutations in *KDM6A* (males 37.5 %, mean age 7.7 years, age range: 0.7 - 17.3 years), and 12 unaffected siblings (males 58.3 %, mean age 14.3 years, age range: 6.3 - 28.8 years). The detailed information of variants, age, data types profiled for each individual is found in **Table 1**. The variant types in *KMT2D* or *KDM6A* are summarized in **Figure 1A**, showing that the majority of them are truncating variants– nonsense, frameshift variants, or deletions (62.5 % for KS1, 87.5 % for KS2).

**Figure 1.**
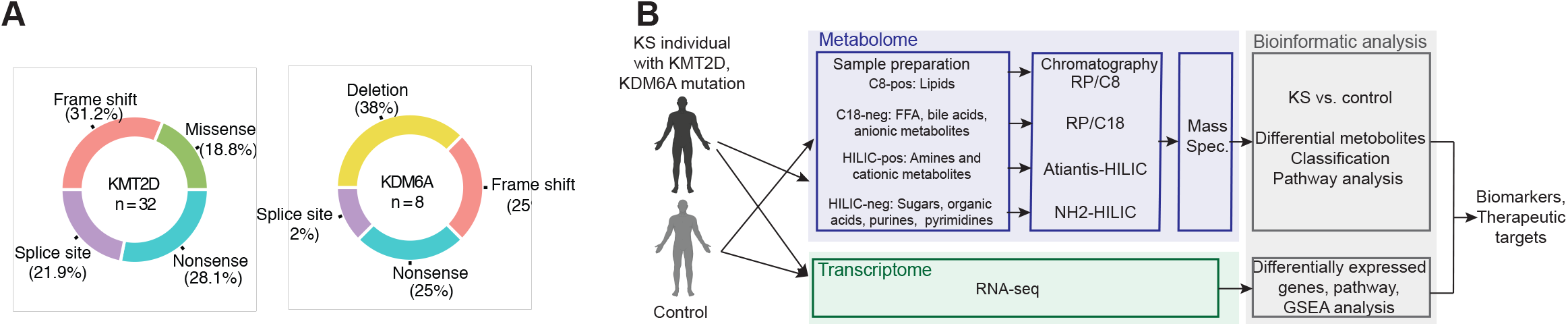
Dataset and variant overview of our cohort. **A.** Proportions of variant types from our KS1 and KS2 cohorts. Red, frame shift; blue, nonsense; green, missense; orange, splice site mutations; yellow, deletions. **B**. Overview of this study design.

The untargeted metabolomics of plasma and urine were profiled for the collected samples (**Figure 1B**). Transcriptomics by RNA-seq of peripheral blood mononuclear cells (PBMCs) were also carried out for a total of 48 subjects among 52 subjects for whom metabolomics was profiled (92.3 %; 29/32 KS1, 8/8 KS2, 11/12 controls).

### Differential metabolites between KS1 and controls

**Since** we have a larger sample size in KS1 (n = 32) compared to KS2 (n = 8), we first focused on KS1 and asked whether the metabolomic landscapes caused by *KMT2D* mutations are altered in KS1. To determine significantly changed metabolites between KS1 groups and control groups, we performed a comparative analysis of the intensities of metabolites identified from plasma and urine samples of KS1 with those of controls. After filtering outliers and normalization of the signals, we statistically accessed the intensity changes between KS and controls, resulting in 151 significantly changed metabolites in KS1 with N2, N2-dimethylguanosine emerging as one of the top metabolites (83 increased, 66 decreased; q=0.1, Student t-test, **Figures 2A and 2B, Table 2**). N2, N2-dimethylguanosine is a urinary metabolomic product, which is a primary product during tRNA degradation. The serum level of this metabolite has been known to be lifted for patients with several cancer types(Tormey et al. 1980; Waalkes et al. 1973; Cho et al. 2009).

**Figure 2.**
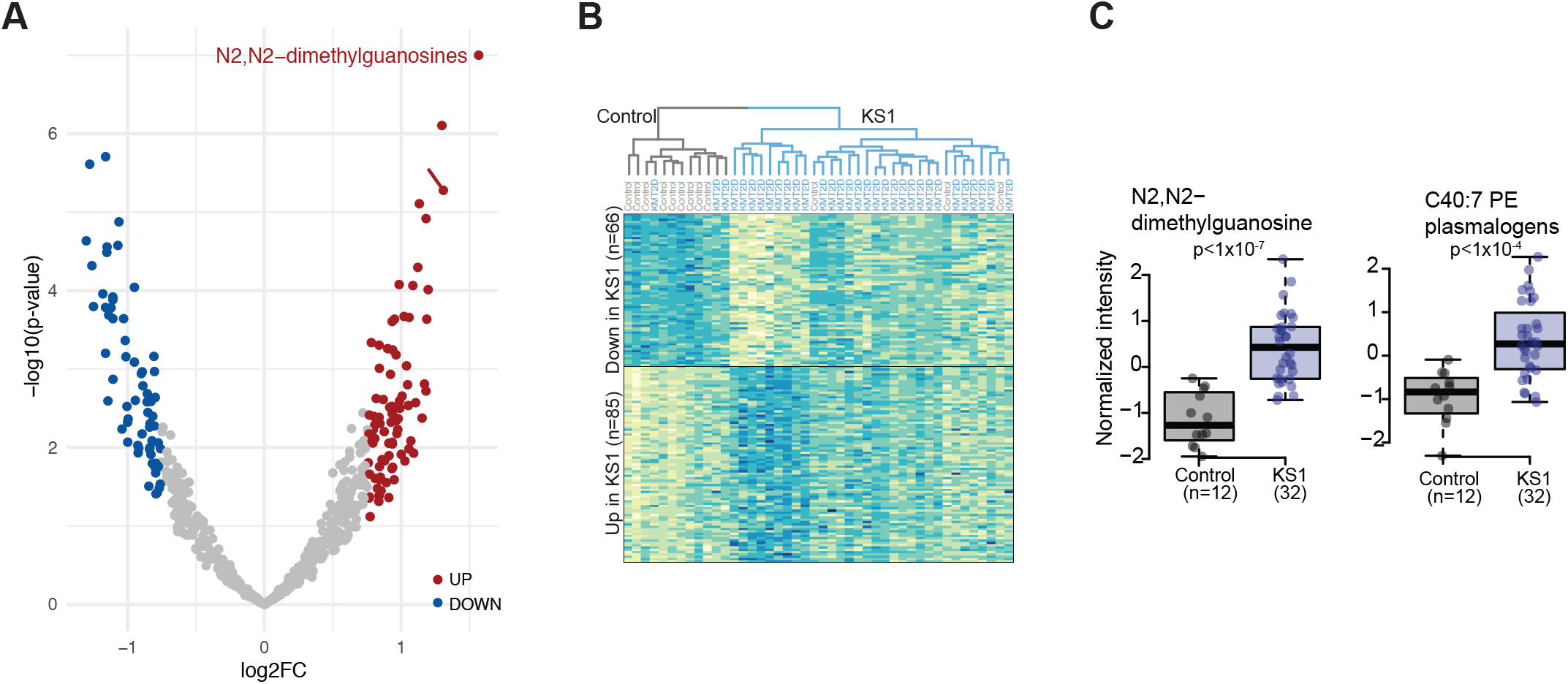
Significantly altered metabolites in KS1. **A.** Volcano plots showing differential metabolites between KS1 and controls. Red: significantly increased (q < 0.1) and blue: significantly decreased (q < 0.1). **B**. Differential metabolites between KS1 and control in heatmap for the normalized metabolite signals. The hierarchical clustering results are shown on the top of the heatmap plot. Blue, high signal intensity; yellow, low signal intensity. **C**. Normalized intensities of the top candidates for KS1 and controls. Each dot represents each sample. P-value by the Student’s t-test.

Hierarchical clustering based on differential metabolites shows the tendency of grouping according to genotypes (**Figure 2B**). The distributions of intensity for the top metabolites are shown for KS1 and controls in **Figure 2C**. This result suggests that the mutations of *KMT2D* affect the metabolomic landscape which is distinctive from that of normal controls.

### Classification between KS1 and controls

Next, we questioned if we can distinguish KS1 from controls using a subset of metabolites. The principle component analysis (PCA) for metabolite signals among individuals revealed that the top ten differential metabolites separated KS1 samples from controls (**Figure 3A**).

**Figure 3.**
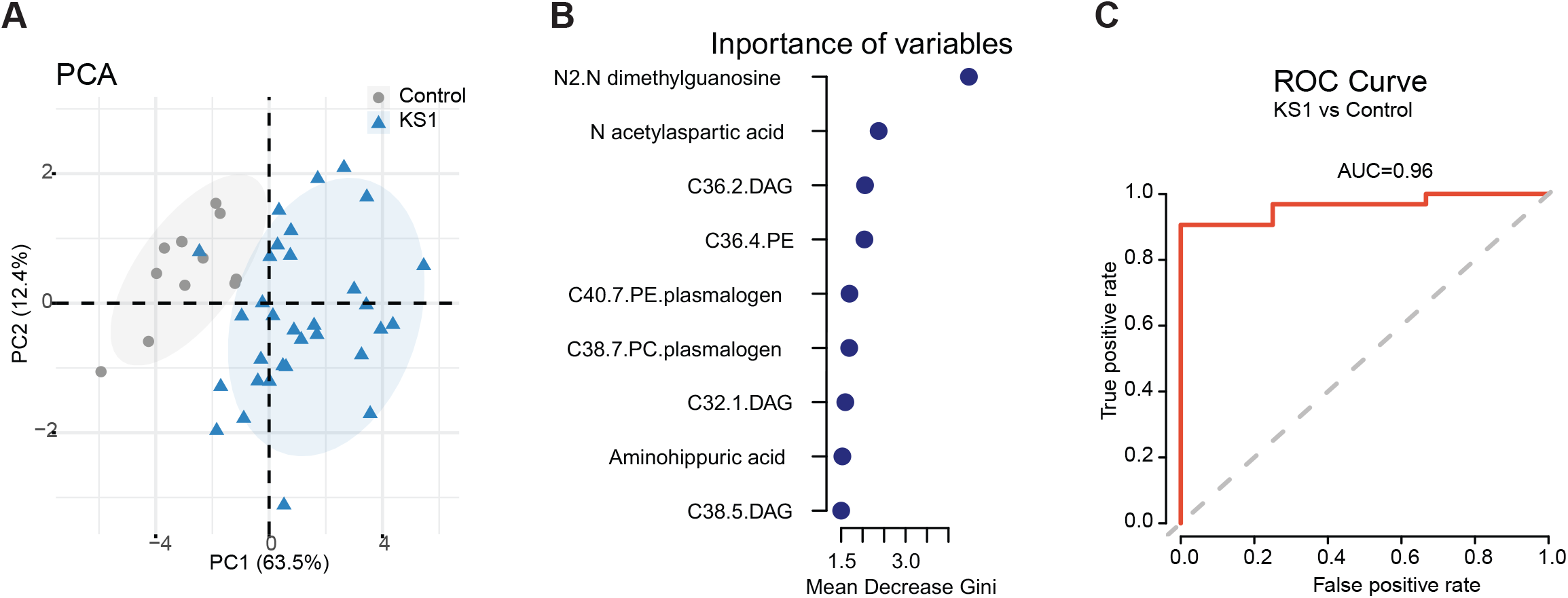
Classification of KS1 and controls. **A.** PCA analysis of the intensity of the top ten metabolites for KS1 and control. Each dot represents an individual sample. Blue, KS1; gray, control. **B**. Importance of metabolites of the Random Forest classifier by the mean decrease of the Gini index. **C**. ROC curve in the Random Forest classifier.

Using machine learning classifiers, Random Forest and Partial Least Squares Discriminant (PLSD), we identified the most crucial metabolites. Once again, N2, N2-dimethylguanosine was determined as the top contributor in both methods (**Figure 3B and Figure S1; Methods**). Our trained models exhibited good performance of distinguishing KS1 from controls (**Figure 3C**, AUC score of 0.96 in Random Forest; R square of 0.92 in PLSD). This suggests that the models based on a set of metabolites could effectively serve as discriminative biomarkers for KS1.

### Pathways associated with altered metabolites in KS1

To examine the metabolomic pathways that are potentially affected by *KMT2D* mutations, we explored the pathways associated with significantly changed metabolites in KS1. We first calculated the significance of enrichment in the KEGG metabolomic pathways for the 151 significantly altered metabolites in KS1 compared to controls.

Additionally, we determined pathway impact scores based on the predicted perturbations in core components for the KEGG metabolomic pathways (**Figure 4A**). The pathways pyrimidine metabolism, alanine, aspartate and glutamate metabolism, pantothenate and CCoA biosynthesis, and sphingolipid metabolism are significantly enriched (p < 0.05) and are expected to be perturbed by altered metabolites in KS1. The top ten pathways with an impact score > 0 are provided in **Table 4**.

**Figure 4.**
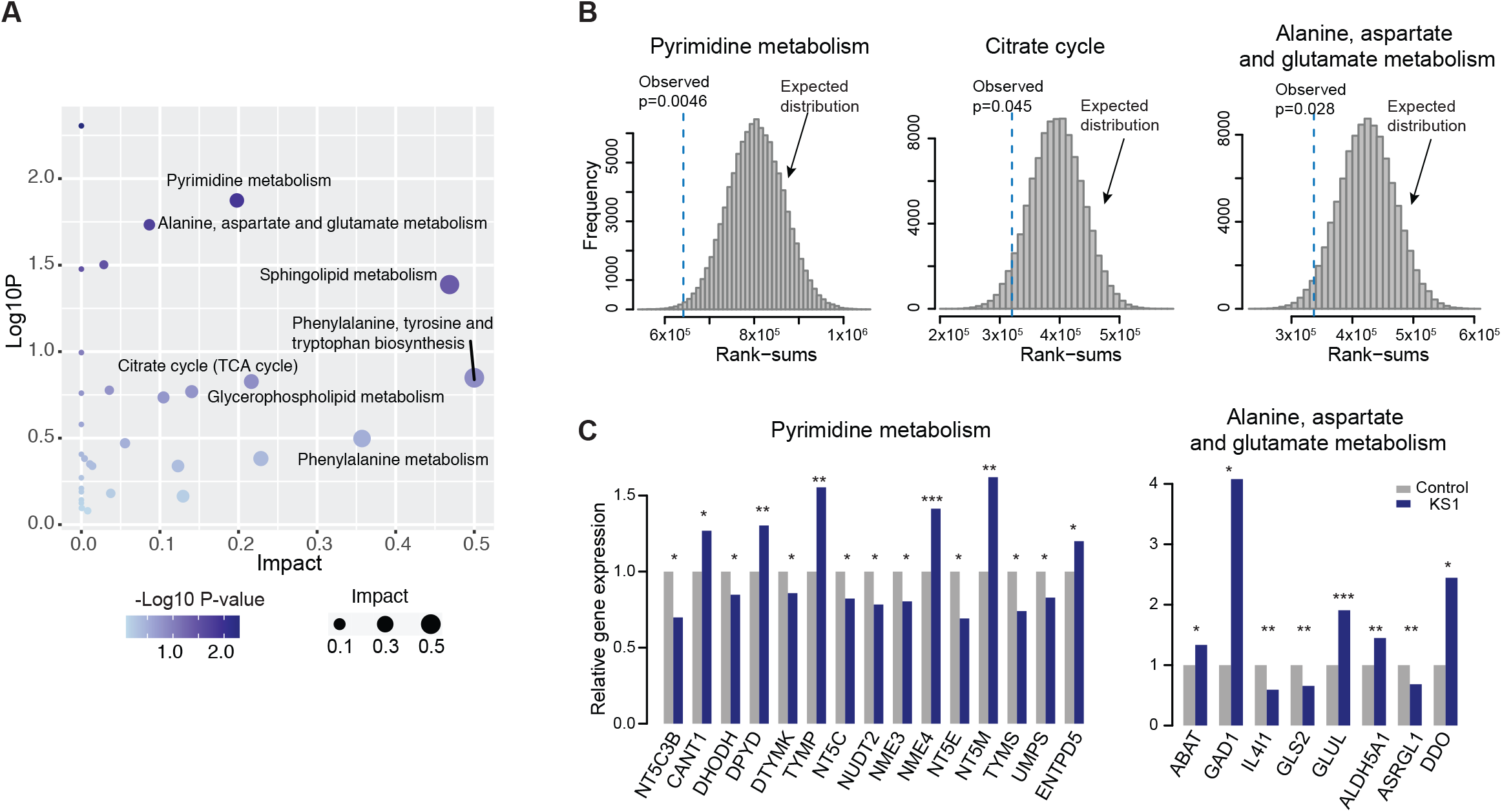
Pathways associated with significantly changed metabolites in KS1. **A.** The enrichment of differential metabolites in KS1 compared to controls for the KEGG metabolic pathways and the predicted impact scores for the pathways. **B**. Expected distribution of the rank-sum statistics and observed one with the p-values based on the rank-sum test for pyrimidine, alanine, citrate cycle, and aspartate and glutamate metabolism pathways. **C**. Expression changes of the genes in pyrimidine, alanine, and aspartate and glutamate metabolism pathways in KS1 compared to control. P-values from a negative binomial model. Blue, KS1; gray, control.

Next, we questioned if the pathways associated with differential metabolites were also disturbed in gene expression in KS1. For the top ten pathways in Table 4, we examined gene expression changes in KS1 compared to controls. Among the ten pathways, three pathways (pyrimidine metabolism, alanine, aspartate and glutamate metabolism, and citrate cycle pathways) exhibit significant gene expression changes compared to the expectation based on background gene expression changes (p < 0.05, rank sum test, **Figure 4B, Methods**). Furthermore, we identified 23 differentially expressed genes between KS1 and controls in pyrimidine metabolism and alanine, aspartate and glutamate metabolism pathways (p < 0.05, a negative binomial model, **Figure 4C**). Notably, several genes with significantly increased expression levels in KS1 have previously been in disease conditions. For instance, TYMP is associated with mitochondrial neurogastrointestinal encephalomyopathy(El-Hattab and Scaglia 2013; J. Wang et al. 2015). GAD1 and DDO have been involved with neurological disorders, including schizophrenia and intellectual disability(Mitchell et al. 2015; Domschke et al. 2013; Cristino et al. 2015; Lombardo et al. 2022). Our results suggest that several metabolomic pathways are perturbed in both the metabolome and transcriptome in KS1, which are associated with pathogenicity in KS.

### Metabolomic changes in KS2

Now shifting our focus to KS2, we examined how the mutations in *KDM6A* affect the metabolomic landscape in KS2. Given the smaller sample size, we identified 10 metabolites that exhibited significant alterations in KS metabolites (8 increased, 2 decreased; q=0.1) compared to those in controls (**Figure 5A**). The ten differential metabolites are shown in **Table 3**. Among the ten differential metabolites in KS2, five were also identified as significantly changed metabolites in KS1. The signal intensities from KS2 for the differential metabolites tend to cluster by the hierarchical clustering analysis (**Figure 5A**). The distributions of intensity for the top metabolites are shown in **Figure 5B for KS2 and controls**. Importantly, N2, N2-dimethylguanosine, which was the top candidate in KS1, appeared as one of the top metabolites in KS2. When comparing the 100 most differential metabolites in KS2 with the 100 most differential metabolites in KS1, we find significant overlap of metabolites between two groups (p = 0.000414, hypergeometric test, **Figure 5C**). Although the ranking of metabolites in KS2 based on significant levels is slightly different from that in KS1, the direction of the fold changes in KS2 compared to controls was consistent with the fold changes in KS1 compared to controls (**Figure S3**). Additionally, we determined the most crucial metabolites for classification between KS2 and controls, using the PLSD method (**Figure S3**). N2, N2-dimethylguanosine was identified as one of the top contributors. The identified metabolites well separated KS2 groups from controls in the PCA analysis (**Figures S3**).

**Figure 5.**
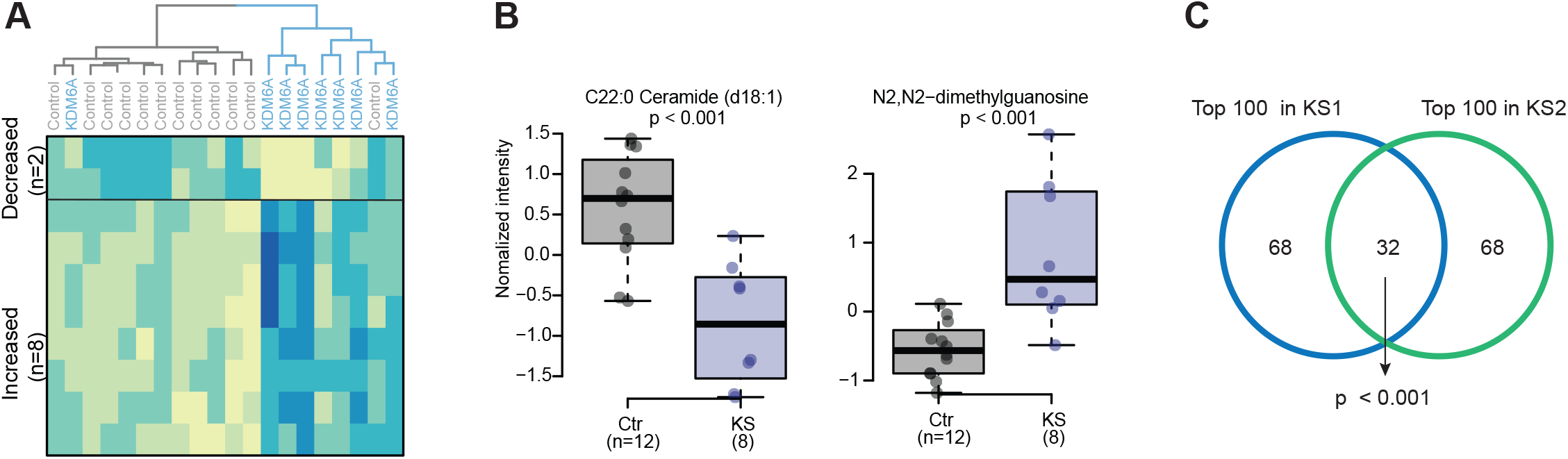
Significantly altered metabolites in KS2. **A.** Differential metabolites between KS2 and control in the heatmap for the normalized metabolite signals. The hierarchical clustering results are shown at the top of the heatmap plot. Blue, high signal intensity; yellow, low signal intensity. **B**. Normalized intensities of the top candidates for KS2 and controls. Each dot represents each sample. P-value by the Student’s t-test. **C**. Overlap between the top 100 differential metabolites between KS1 and control and top 100 differential metabolites between KS2 and control. P-value by the hypergeometric test.

Next, we examined the metabolomic pathways that might be affected in KS2 caused by *KDM6A* mutations. Due to the limited sample size in KS2 samples, we used a relaxed threshold of p-value of 0.05 instead of q-value of 0.1, resulting in 118 differential metabolites in KS2. For these metabolites, we assessed the enrichment in the KEGG metabolomic pathways and calculated impact scores for the prediction of pathway perturbations by the altered metabolites (**Figure S3**). Alanine, aspartate and glutamate metabolism, Pyrimidine metabolism, arginine biosynthesis, and sphingolipid metabolism pathways are significantly enriched (p < 0.05) and are expected to be perturbed by altered metabolites in KS2. Importantly, among the four pathways, three of them (alanine, aspartate and glutamate metabolism, pyrimidine metabolism, and sphingolipid metabolism pathways) were also identified as significantly enriched pathways in KS1. The top ten pathways with an impact score > 0 are provided in **Table 5**. Taken together, these results suggest that a significant number of metabolites exhibited changes in the same direction in both KS1 and KS2. The similar pathways are likely to be affected by these metabolite alterations in both KS1 and KS2.

## DISCUSSION

To examine the alterations in the metabolomic landscape in KS, we profiled the untargeted metabolome from urine and plasma samples of 40 KS patients with pathogenic variants in *KMT2D* or *KDM6A* together with transcription data and systematically identified altered metabolites and associated pathways in KS.

Although there was genetic heterogeneity in individuals with KS1 and KS2, we detected a large number of the same metabolites that underwent significant changes in both KS1 and KS2. Furthermore, similar pathways tended to be affected by the metabolite alterations in both KS1 and KS2. This suggests that although there are differences in the mechanisms caused by mutations in *KMT2D* and *KDM6A*, their targets and effects at the metabolomic level are similar on a large scale. However, we also observed differences on a smaller scale, which requires a systematic comparative study using data from a larger cohort.

N2, N2-dimethylguanosine was significantly elevated both in KS1 and KS2 compared to controls. The level of this metabolite has been reported to increase in patients with several cancers(Tormey et al. 1980; Waalkes et al. 1973; Cho et al. 2009). Previous studies on cancers and KS models with *KMT2D* or *KDM6A* mutations suggested that the condition causes activation of several pathways including Pyrimidin and RAS/MAPK pathways(Pan et al. 2023; Lu et al. 2023; Tsai et al. 2018). Therefore, the elevated levels of N2, N2-dimethylguanosine might be associated with common pathways between KS and cancers.

It is pertinent to note that the comparison analysis performed was limited between KS and controls in this study. It has been noted that individuals with various developmental disorders, in particular sharing mechanisms such as mutations in chromatin modifiers, present with similar phenotypes as in Kabuki syndrome (Gabriele et al. 2018; Ronan et al. 2013; Fahrner and Bjornsson 2014). The common symptoms make it challenging to precise diagnosis at earlier ages. Although this study suggests a set of metabolites that distinguishes KS from controls, it is not clear which metabolites behave differently between KS and other developmental disorders. However, it is likely that this strategy using differential metabolites for biomarkers could be applied for other disorders. The future expanding to other developmental disorders will allow us to better understand insights into disorder-specific metabolomic landscapes.

In conclusion, although there was genetic heterogeneity in individuals with KS1 and KS2, we identified metabolites in plasma and urine that distinguished KS from controls, suggesting that the differential metabolites could serve effective biomarkers for KS. In particular, N2, N2-dimethylguanosine emerged as one of the top candidates both for KS1 and KS2, holding potential as a promising biomarker.

## METHODS

### Data collection

Plasma and urine samples were collected from children clinically diagnosed with KS, confirmed by genetic testing with mutations in either *KMT2D* or *KDM6A* through the Roya Kabuki Program at Boston Children’s Hospital. Enrolled individuals with Kabuki syndrome and their family members provided consent for research participation. Additionally, samples from unaffected parents or siblings of the patients were collected, when available.

### Metabolomic profiling

Four complementary liquid chromatography tandem mass spectrometry (LC-MS) methods were employed to quantify lipids and polar metabolites in plasma and urine samples, as previously described (T. J. Wang et al. 2013; Mascanfroni et al. 2015; Esko et al. 2017; Paynter et al. 2018; Lloyd-Price et al. 2019). The methods are characterized by the chromatography stationary phase and MS ionization mode used and are referred to as C8-pos, C18-neg, HILIC-pos, and HILIC-neg. The C8-pos, C18-neg, and HILIC-pos methods were configured on LC-MS systems comprised of Nexera X2 U-HPLCs (Shimadzu) coupled to Q Exactive series orbitrap mass spectrometers (Thermo Fisher Scientific) for high resolution accurate mass (HRAM) profiling of both hundreds of identified metabolites and thousands of unknowns, while the HILIC-neg method was operated on both a Nexera X2-Q Exactive system for HRAM profiling and a UPLC (Waters) coupled to a QTRAP 5500 (SCIEX) for targeted profiling.

The C8-pos method measures polar and nonpolar lipids. Lipids were extracted from 10 μL plasma using 190 μL of isopropanol, separated using reversed phase C8 chromatography, and analyzed HRAM, full-scan MS in the positive ion mode. The C18-neg method measures free fatty acids, oxidized fatty acids and lipid mediators, bile acids, and metabolites of intermediate polarity; these metabolites were extracted from 30 μL plasma using 90 μL of methanol, then separated using reversed phase C18 chromatography, and analyzed using HRAM, full-scan MS in the negative ion mode. The HILIC-pos method measures amino acids, amino acid metabolites, acylcarnitines, dipeptides, and other cationic polar metabolites; these metabolites were extracted from 10 μL plasma using 90 μL of 25% methanol/75% acetonitrile, then separated using hydrophilic interaction liquid chromatography (HILIC), and analyzed using HRAM, full-scan MS in the positive ion mode. The HILIC-neg method measures sugars, organic acids, purines, pyrimidines, and other anionic polar metabolites; these metabolites were extracted from 30 μL plasma using 120 μL of methanol containing internal standards and analyzed using either HRAM, full-scan MS in the negative ion mode or targeted multiple reaction monitoring using the QTRAP 5500 triple quadruple MS system.

Raw data from Q Exactive series mass spectrometers were processed using TraceFinder software (Thermo Fisher Scientific) to detect and integrate as subset of identified metabolites and Progenesis QI software (v 2.0, Nonlinear Dynamics) to detect, de-isotope, and integrate peak areas from both identified and unknown metabolites. MultiQuant (SCIEX) was used to integrate peak areas of metabolites measured using the QTRAP 5500. Identities of metabolites were confirmed by matching measured retention times (RT) and mass-to-charge ratios (m/z) to authenticate reference standards. Unnamed metabolites were not used in the downstream analyses.

### Transcriptomic profiling

RNA-seq data was obtained for transcriptomic profiling, as described before(Jung et al. 2023). Total RNA was extracted from whole blood following the manufacturer’s protocol for PAXgene Blood RNA Kit (Qiagen). Following the assessment of RNA concentration and quality (RIN > 7.2) by Agilent 2100 or Fragment Analyzer, the illumina TruSeq Stranded Total RNA with Ribo-Zero Globin library preparation was used, which includes ribosomal RNA and globin mRNA removal. A minimum of 10Gb of 100bp paired-end reads were generated per sample. All library preparations and sequencing was conducted by BGI Americas on the illumina HiSeq 4000 platform.

### Metabolomic analysis

For peak annotation, the detected peaks were annotated with putative compound identities using reference MS/MS spectra libraries. Peak intensities were normalized by calculating the median, followed by a log-transformation. Subsequently, they were standardized to generate z-scores.

For statistical analysis, normalized peak intensities were compared between KS groups and control groups. Significance were determined using statistical tests, the Wilcoxon rank-sum test or the Student’s t-test. Adjustments for multiple comparisons were applied.

To classify KS and control groups, we employed a Random Forest and a multivariate regression model(the partial least squares discriminant analysis, PLS-DA). The metabolites that are most discriminative between KS and control groups were identified.

For pathway analysis, MetaboliteAnalysis was used to determine the enrichment and the pathway impact scores in KEGG metabolomic pathways. Disrupted metabolic pathways were associated with the significantly altered metabolites in KS.

### Transcriptomic analysis

The RNA-seq data were integrated with the metabolomic data to gain insights into the affected pathways. For the KEGG metabolic pathways associated with altered metabolites in KS, the gene expression levels from RNA-seq data were compared between KS and controls to assess the significance using DESeq2. Age and sex were taken into account as covariates in the statistical modeling.

The results were visualized using various techniques, including Principal Component Analysis (PCA), heatmaps, hierarchical clustering, and ROC curve plots.

## Data Availability

All data produced in the present study are available upon reasonable request to the authors

## ACKNODGEMENTS

This study was funded by the Philanthropic Fund for the Roya Kabuki Program. YLJ was supported by the Boston Children’s Hospital Office of Faculty Development**/**Basic & Clinical Translational Research Executive Committees Faculty Career Development Fellowship.

## FIGURE LEGENDS

**Table 1.** Summary of patients, variants, metabolome, and transcriptome

**Table 2.** Top ten differential metabolites in KS1 compared to controls

**Table 3.** Top ten differential metabolites in KS2 compared to controls

**Table 4.** Top ten pathways associated with differential metabolites in KS1

**Table 5.** Top ten pathways associated with differential metabolites in KS2

